# Hybrid Neural–Bayesian Belief Network Framework for Uncertainty-Aware Multimodal GBM Prediction

**DOI:** 10.64898/2026.05.10.26352710

**Authors:** Alejandra Jayme, Vincent Heuveline

**Affiliations:** Independent Researcher, Berlin, Germany; Ruprecht-Karls -Universität Heidelberg, Heidelberg, Germany

**Keywords:** Bayesian neural networks, Bayesian belief networks, radiogenomics, glioblastoma, uncertainty quantification, multimodal learning

## Abstract

**Background and Objective:** Glioblastoma outcome prediction remains difficult because clinically relevant signals are distributed across heterogeneous imaging and genomic modalities, cohorts are small, and conventional neural predictors do not quantify their own uncertainty. This study evaluates a hybrid neural–Bayesian belief network framework for uncertainty-aware multimodal glioblastoma prediction and examines how modality selection, model family, and structure-aware regularization affect predictive performance and confidence quality.

**Methods:** The framework was evaluated on the TCGA-GBM radiogenomic cohort using four input modalities (T1Gd, FLAIR, mRNA, and CNA), five model families, five structural-weight settings, and 15 view subsets. A secondary benchmark on the UCI Human Activity Recognition dataset was included to assess whether observed limitations were specific to the glioblastoma setting.

**Results:** CNA features consistently reduced performance in most multimodal settings, and selective fusion excluding CNA outperformed both the full four-view baseline and imaging-only alternatives. Model families showed clear differences in uncertainty behaviour: non-Bayesian families achieved the strongest predictive accuracy, whereas the Bayesian family achieved the lowest calibration error over a narrower confidence range. Bayesian belief network regularization produced consistent directional improvements without supporting reliable structure-discovery claims, as learned graph structures were not reproducible across folds. On the secondary benchmark, the same framework achieved much higher predictive performance, indicating that the glioblastoma performance ceiling primarily reflects data limitations rather than an architectural constraint.

**Conclusions:** In small-sample radiogenomic prediction, modality choice is at least as important as model choice, and uncertainty quality differs substantially across uncertainty-aware model families. The proposed framework provides a practical basis for comparing accuracy, calibration, modality selection, and structure-aware regularization in multimodal biomedical prediction.

## 1 Introduction

Glioblastoma (GBM) remains the most aggressive primary brain tumor in adults, with outcomes remaining poor despite multimodal treatment [2, 3]. This makes prognosis and patient stratification important, but also difficult: the disease is heterogeneous, the available cohorts are limited in size, and clinically relevant signals are distributed across different data types rather than concentrated in any single modality. Predictive modeling is therefore attractive in GBM not only as a classification exercise but as a way to combine heterogeneous evidence into a more useful estimate of outcome risk.

Radiogenomics is especially promising in this setting because it links non-invasive imaging with molecular information [4]. Magnetic resonance imaging (MRI) captures spatially structured macroscopic phenotypes, whereas genomic measurements capture molecular mechanisms that may underlie aggressiveness, treatment response, and survival. However, these modalities are not naturally matched in scale or statistical structure. Imaging features are relatively low dimensional and organized around anatomy and texture, while genomic features are high dimensional, sparse, and often noisy. Building predictive models that use both well is therefore a nontrivial multimodal learning problem.

Deep learning offers a natural route for representation learning across modalities, yet two well-known obstacles remain central in clinical use. First, standard neural predictors often return point estimates without a meaningful notion of predictive confidence. In small-sample biomedical settings, this is a serious limitation: uncertainty may arise from cohort heterogeneity, data noise, and model misspecification, and ignoring it can lead to unwarranted trust in incorrect predictions. Second, even when predictive performance is acceptable, the internal reasoning of a deep model is usually opaque. That opacity complicates scientific interpretation and reduces clinical trust [5].

Bayesian deep learning is attractive because it addresses at least part of this gap. By introducing stochasticity or probabilistic weight modeling, Bayesian or Bayesian-inspired models can represent predictive uncertainty more explicitly than deterministic baselines [6, 7]. At the same time, probabilistic graphical models can encode conditional dependencies in a form that is more interpretable than raw neural activations alone. These ideas motivate hybrid approaches that aim to combine representation learning, uncertainty quantification, and structured reasoning.

This paper studies such a hybrid Bayesian deep learning framework, combining neural prediction with a Bayesian belief network (BBN) component. The BBN component functions here as a structure-aware regularizer and component diagnostic rather than a validated graph-recovery method. Instead of treating uncertainty-aware modeling as a single method choice, we instantiate the framework as a structured family of deterministic, heuristic uncertainty-aware, and variational Bayesian variants.

We evaluate the framework as a multimodal GBM prediction system and ask the following questions:

- Which model family performs best on multiview GBM prediction?
- How do calibration, overconfidence, and epistemic uncertainty differ across model families?
- How does the structural regularization weight affect predictive behavior?
- Which imaging, genomic, and multimodal views contribute most to predictive performance?

The contribution of this paper is a systematic comparative analysis that yields three concrete findings. First, selective multimodal fusion (combining T1Gd, FLAIR, and mRNA) outperforms naive four-view fusion across the non-Bayesian model families; adding CNA consistently degrades accuracy across most contexts. Second, predictive accuracy, calibration, and epistemic uncertainty trade off across families: non-Bayesian families achieve the highest accuracy, the Bayesian family achieves the lowest ECE over a narrower confidence range, and epistemic uncertainty signals differ across families. Third, BBN structural regularization acts as a latent regularizer rather than a structure-recovery mechanism: it improves accuracy and calibration consistently, but learned graph structures are fold-dependent and not reproducible. Together these results clarify which uncertainty strategies are most effective under realistic small-cohort multimodal conditions.

## 2 Related Work

### 2.1 GBM and Radiogenomic Prediction

Prior work in glioblastoma has established both the clinical urgency of prognosis modeling and the value of integrated radiogenomic resources for studying tumor heterogeneity [2, 3, 8–11]. More broadly, radiogenomics has been framed as a way of linking imaging phenotypes with molecular information for diagnosis, prognosis, and treatment stratification across cancer settings [4]. At the same time, radiogenomic prediction remains methodologically difficult because imaging and molecular measurements differ substantially in scale, noise structure, and biological granularity. These practical constraints motivate comparative multimodal studies rather than single-model claims based on one chosen fusion strategy.

Specific GBM studies have used radiogenomic pipelines to predict molecular features and survival-related outcomes from MRI-derived features. Machine-learning analyses combining MRI radiomics with metagene information have stratified GBM patients by survival group [12], while uncertainty-aware radiogenomic modeling has been explored for region-level prediction of EGFR amplification [13]. More recent work has integrated multiparametric MRI texture features with molecular biomarkers for survival classification [14], and multimodal deep learning combining histopathology with gene expression has demonstrated consistent prognosis improvement over unimodal models across adult and pediatric brain tumors, with early fusion identified as the most robust integration strategy [15]. Gomaa et al. developed a transformer-based multimodal glioblastoma survival model integrating MRI with clinical and molecular-pathologic inputs across multicenter cohorts [16].

While many studies focus on optimizing individual radiogenomic pipelines, this work positions itself as a framework paper for multimodal GBM prediction under uncertainty, with particular emphasis on view selection and calibration rather than just raw predictive performance.

### 2.2 Uncertainty-Aware and Bayesian Medical AI

Uncertainty-aware modeling has become increasingly important in medical AI because predictive confidence matters in high-stakes settings. Bayesian deep learning and approximate Bayesian methods such as Monte Carlo dropout have been proposed as practical ways to represent predictive uncertainty in neural models [6, 7]. More broadly, uncertainty and interpretability are often discussed together because both address the mismatch between high-performing black-box systems and the needs of clinical decision support [5]. In the GBM radiogenomics setting, uncertainty-aware modeling has already been argued to improve the reliability and interpretability of molecular prediction, even when predictive performance remains sensitive to data regime and task formulation [13]. Bayesian neural networks have also been applied directly to survival prediction, where credible intervals over predicted survival distributions provide more informative uncertainty estimates than point predictions alone [17]. At the level of model evaluation, calibration quality has emerged as a distinct axis of comparison from discrimination performance, with uncertainty-aware training shown to reduce overconfidence in clinical deep learning settings [18].

This work differs from a pure uncertainty-method paper in two ways. First, it studies uncertainty within a clinically motivated multimodal GBM setting rather than in a generic benchmark-only context. Second, it compares deterministic, heuristic uncertainty-aware, and fully probabilistic variants within a single pipeline, allowing practical trade-offs to be evaluated directly rather than inferred from separate studies.

### 2.3 Multimodal and Structured Probabilistic Modeling

Multimodal medical prediction models combine imaging and molecular measurements, but such models often remain either purely discriminative or only weakly connected to explicit probabilistic structure. Large-scale efforts such as PORPOISE [19] have demonstrated structured multimodal fusion of histopathology and multi-omics data across thousands of patients, while VAMBN [20] combined modular variational autoencoders with a Bayesian network over the latent space to model dependencies between heterogeneous clinical variable groups. VAMBN is the closest architectural prior to the neural network–belief network hybrid studied here. In parallel, probabilistic graphical modeling remains attractive because it provides structured conditional dependencies that are easier to inspect than raw neural activations.

This works sits between these traditions: a neural latent representation carries the predictive burden, while a Bayesian network regularization component introduces an explicit structural constraint over latent variables. The aim is to study whether a hybrid framework combining these two components changes predictive behavior, uncertainty behavior, and multimodal value in a practically meaningful way. The distinctive emphasis relative to deterministic multimodal predictors [15, 16] is not only on multimodal prediction strength, but on calibration, epistemic uncertainty, leave-one-view-out contribution, and the effect of structured probabilistic regularization within a single comparative pipeline.

## 3 Methods

### 3.1 Datasets

#### GBM cohort

The primary dataset is the TCGA-GBM radiogenomic cohort. Clinical and genomic data are drawn from the TCGA GBM Firehose Legacy study [8, 10, 11]; imaging features are taken from the BraTS-TCGA-GBM collection on TCIA [9, 21]. The cohort comprises 134 patients with complete imaging data; genomic modalities have lower coverage due to assay availability (124 with mRNA, 129 with CNA). For multiview combinations, a patient is included if and only if they have complete data for every constituent modality; the resulting intersection counts are shown in Table 1. The prediction target is a three-class overall-survival label (OS_LABEL) derived from OS_MONTHS using fixed thresholds: class 0 (short, ≤ 8.4 months, *n* = 45), class 1 (mid, 8.4–18.5 months, *n* = 45), and class 2 (long, *>* 18.5 months, *n* = 44). Thresholds are fixed at cohort construction and are not refit per fold.

**Table 1:** GBM cohort summary. Feature counts are before per-fold Cox selection (10 selected per view per fold). Overlap rows show the patient intersection and total raw features for each view combination.

| Modality | Feature type | Patients | Raw features |
| --- | --- | --- | --- |
| T1Gd radiomics | Volume, shape, TGM, intensity, texture (ET/ED/NET) | 134 | 229 |
| FLAIR radiomics | Volume, shape, TGM, intensity, texture (ET/ED/NET) | 134 | 229 |
| mRNA | Gene expression OncoKB ∩ mutated | 124 | 199 |
| CNA | Copy-number OncoKB genes | 129 | 396 |
| *Two-view combinations* |  |  |  |
| t1gd_flair | Imaging only | 134 | 458 |
| t1gd_mrna | T1Gd + mRNA | 124 | 428 |
| t1gd_cna | T1Gd + CNA | 129 | 625 |
| flair_mrna | FLAIR + mRNA | 124 | 428 |
| flair_cna | FLAIR + CNA | 129 | 625 |
| mrna_cna | Genomic only | 119 | 595 |
| *Three-view combinations* |  |  |  |
| t1gd_flair_mrna | Imaging + mRNA | 124 | 657 |
| t1gd_flair_cna | Imaging + CNA | 129 | 854 |
| t1gd_mrna_cna | T1Gd + genomic | 119 | 824 |
| flair_mrna_cna | FLAIR + genomic | 119 | 824 |
| *Four-view combination* |  |  |  |
| t1gd_flair_mrna_cna | Full multimodal | 119 | 1,053 |
| *OS_LABEL class distribution ($n = 134$)* |  |  |  |
| Class 0 | Short survival ($\leq 8.4$ months) | 45 | |
| Class 1 | Mid survival (8.4–18.5 months) | 45 |  |
| Class 2 | Long survival ($> 18.5$ months) | 44 | |

Four modalities are used. Imaging features for T1Gd and FLAIR sequences are taken from the pre-computed BraTS-TCGA-GBM radiomics tables [21], which provide 229 features per MRI sequence computed across three BraTS tumor sub-regions: enhancing tumour (ET), peritumoral oedema (ED), and non-enhancing tumour/necrosis (NET). Feature groups cover volumetric ratios and absolute volumes, spatial location, shape descriptors (eccentricity, solidity), Tumour Growth Model (TGM) parameters, intensity statistics, 10-bin histograms, and four texture matrix families (GLCM, GLRLM, GLSZM, NGTDM) [22]. Shared non-sequence features (volumes, shape, TGM) are included in each sequence view. Genomic features are filtered from TCGA GBM Firehose Legacy RNA-seq and SNP6 array data using two gene-set profiles. For mRNA, features are restricted to the intersection of OncoKB cancer genes [23] and genes recurrently mutated in GBM, yielding 199 genes. For CNA, features are restricted to OncoKB cancer genes regardless of GISTIC significance [24] or alteration frequency, yielding 396 genes. Table 1 summarises per-modality counts and multiview overlaps.

#### Feature selection and preprocessing

Within each training fold, features are selected independently per view using univariate Cox partial-likelihood ranking: each candidate is scored by the gain in Breslow partial log-likelihood when used as a single covariate, with fold-local OS_MONTHS and OS_STATUS as the survival endpoint. The top 10 features per view are retained, enforcing equal input dimensionality across modalities regardless of their intrinsic informativeness; this was a pragmatic design choice rather than a principled claim. Selected features are standardised with a StandardScaler fitted on the training fold only; the same scaler is applied to the test fold without refitting.

#### HAR benchmark

The UCI HAR dataset is included as a secondary benchmark to test whether model-ranking and uncertainty patterns generalise beyond small clinical data [25]. The corpus contains 10,299 windows of smartphone inertial sensor recordings from 30 volunteers performing six activities (WALKING, WALKING_UPSTAIRS, WALKING_DOWNSTAIRS, SITTING, STAND-

ING, LAYING), preprocessed into 561 hand-engineered time- and frequency-domain features. This study partitions those features into four named views by signal type (Table 2), discards the canonical train/test split, and applies the same five-fold stratified cross-validation as GBM over the full 10,299-window pool. Keeping *k* = 5 fixed across datasets was an implementation decision made for protocol consistency and computational comparability with the GBM experiments, rather than a claim that five folds are uniquely optimal for HAR. Within each training fold, the top 50 features per view are selected by Median Absolute Deviation (MAD) and standardised with a fold-local `StandardScaler`. The primary HAR reporting view `t_acc_t_gyro` (time-domain accelerometer and gyroscope) was designated post-hoc after running the HAR view sweep; results for this view are reported in Section 4.6.

**Table 2:** HAR view partition and multiview combinations. Raw feature counts are before per-fold MAD selection (50 features per base view are used per fold).

| View | Signal group | Domain | Raw features |
| --- | --- | --- | --- |
| **t_acc** | Body/gravity accelerometer, jerk, magnitudes | Time | 159 |
| **t_gyro** | Body gyroscope, jerk, magnitudes | Time | 106 |
| **f_acc** | Body accelerometer, jerk, magnitudes | Frequency | 184 |
| **f_gyro** | Body gyroscope, magnitudes | Frequency | 105 |
| *Multiview combinations* |  |  |  |
| **t_acc_t_gyro** | Time-domain accelerometer + gyroscope (primary) | Time | 265 |
| **f_acc_f_gyro** | Frequency-domain accelerometer + gyroscope | Frequency | 289 |
| *Activity class distribution ($n = 10,299$)* |  |  |  |
| Class 0 | WALKING | 1,722 |  |
| Class 1 | WALKING_UPSTAIRS | 1,544 |  |
| Class 2 | WALKING_DOWNSTAIRS | 1,406 |  |
| Class 3 | SITTING | 1,777 |  |
| Class 4 | STANDING | 1,906 |  |
| Class 5 | LAYING | 1,944 |  |

### 3.2 Feature Views and Cross-Validation

For GBM, the experiment uses a structured set of single-view and multimodal combinations rather than only one fused input. This supports two different goals at once: a full-view comparison for the main predictive analysis, and a per-view contribution analysis for understanding which modalities carry most of the predictive signal.

The experiment covers 15 views comprised of all 4 single-view, 6 two-view, 4 three-view, and 1 four-view combination of the four modalities. The full four-view combination serves as the reference for leave-one-view-out analysis. Together, the 15 views support single-view baselines, pairwise comparisons, and a complete marginal contribution analysis at every level of combination. The designation of `t1gd_flair_mrna` as the primary view for the main predictive comparison is *post-hoc*: all 15 views were run first, and the three-view imaging–mRNA combination was selected as the focus view after the view sweep identified it as the highest-accuracy combination (Section 4.5). The *β* sweep, uncertainty, and structural weight analyses are therefore conducted on this post-hoc selected view. This selection is not blind and the specific view’s results should not be interpreted as an independent confirmation; the view analysis in Section 4.5 provides the broader context across all combinations.

All primary analyses are based on five-fold cross-validation. Model selection is not done by assuming a best structural-weight value in advance. The best-performing *β* is selected per model from the observed sweep, keeping the comparisons empirically grounded.

### 3.3 Model Families and Architecture

All model families share a common multimodal encoder backbone and differ only in how uncertainty is represented in the output path. The backbone is identical across families: each input view is processed by a separate view subnet, the resulting view latents are concatenated and projected through a shared cross-view fusion layer, and the resulting latent representation **z** is passed to the output head.

**View subnets.** Each view subnet is a two-layer MLP with the compact architecture:

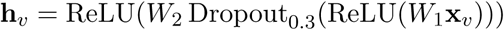

where *W*_1_ ∈ R^32×^*^dv^* and *W*_2_ ∈ R*^d^*^lat×32^, *d_v_* is the number of selected features for view *v*, and *d*_lat_ = ⌊*d_v_/*2⌋ is the per-view latent dimension (e.g. *d*_lat_ = 5 for the GBM primary view with 10 selected features per view; this halving rule was a practical default and was not optimised). The first-layer weight matrix *W*_1_ is used post hoc for feature-to-latent contribution analysis.

**Fusion layer.** View latents are concatenated and projected to the shared cross-view latent space:

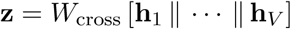

where *W*_cross_ ∈ R*^dz^* ^×(^*^V^* ^·^*^d^*^lat)^ and *d_z_* = *V* · *d*_lat_ is the cross-view latent dimension (*d_z_* = 15 for the three-view GBM primary view; *d_z_* = 50 for the two-view HAR primary view). This **z** is the canonical interface between the encoder and both the output head and the BBN structural component. Critically, **z** is always the pre-dropout, deterministic projection from *W*_cross_. Dropout or stochastic sampling in the output head operates on a separate copy **z̃** = Dropout_0.3_(**z**), leaving **z** unmodified. This design ensures the structural component always receives the canonical latent representation regardless of which uncertainty mechanism is active in the output path.

**Output head per model family.** The five model families differ only in the form of the prediction head and the uncertainty mechanism applied to the shared latent representation **z**.

- `nn`: deterministic. **ŷ** = *W*_out_ **z**. Dropout in view subnets is disabled at test time; no stochasticity in the forward pass.
- `ll` (local likelihood): stochastic output head, no dropout on **z**. Two parallel linear heads predict ***µ*** = *W_µ_* **z** and log ***σ*** = *W_σ_* **z**; logits are sampled as **ŷ** ∼ N (***µ****, e*^log^ ***^σ^***) and converted to class probabilities via softmax. Aleatoric uncertainty is captured by ***σ***.
- `mc_dropout`: dropout on **z̃**, deterministic output head. **ŷ** = *W*_out_ **z̃**. At test time, *T* = 100 stochastic forward passes with dropout active approximate the predictive distribution [6].
- `mc_dropout_ll`: dropout on **z̃**, stochastic output head. **ŷ** ∼ N (*W_µ_* **z̃***, e^Wσ^* **^z̃^**). Combines epistemic uncertainty from dropout with aleatoric uncertainty from the stochastic head. *T* = 100 test-time passes.
- bayesian: fully probabilistic variational inference via Pyro [26]. The layer weights are treated as random variables with a Normal priors; a mean-field Normal guide is fit by ELBO optimisation using the AutoNormal approximation [27]. Predictive uncertainty is obtained by averaging *T* = 100 posterior samples.

**Dropout placement.** Two distinct dropout mechanisms are present in the architecture. The view-subnet Dropout_0.3_ (see View subnets above) is a regularization dropout applied inside every view encoder during training only; it is disabled at test time for all five model families. The `mc_dropout` and `mc_dropout_ll` families apply a separate Dropout_0.3_ to the fused latent **z** to produce **z̃** (see Fusion layer above); this dropout remains active at test time and is the sole source of epistemic uncertainty in those families. Uncertainty estimates for `mc_dropout` and `mc_dropout_ll` therefore arise from the post-fusion dropout, not the in-subnet regularization dropout. The nn, ll, and bayesian families have no dropout active at test time.

This grouping covers three practical uncertainty categories: deterministic prediction (nn), heuristic uncertainty-aware prediction (ll, `mc_dropout`, `mc_dropout_ll`), and fully probabilistic Bayesian inference (bayesian).

### 3.4 Hybrid Framework

The hybrid framework combines a neural network (NN) predictor with a Bayesian belief network (BBN) over latent variables and the outcome. The same multimodal encoder is shared across all model families; the families differ only in the output path, as described above. The BBN consists of a directed acyclic graph (DAG) learner and a Structure Network (SN), which together define the structural regularization mechanism. The latent representation **z** produced by the fusion layer is used as the interface between high-dimensional multimodal inputs and the probabilistic structure model. During training, a predictive objective is augmented with a structure-related regularization term weighted by *β*; when *beta* = 0 the BBN branch is inactive. This design allows the neural component to remain the main predictive engine while encouraging latent representations that are compatible with structured probabilistic dependencies. Figure 1 summarizes the architecture.

**Figure 1:**
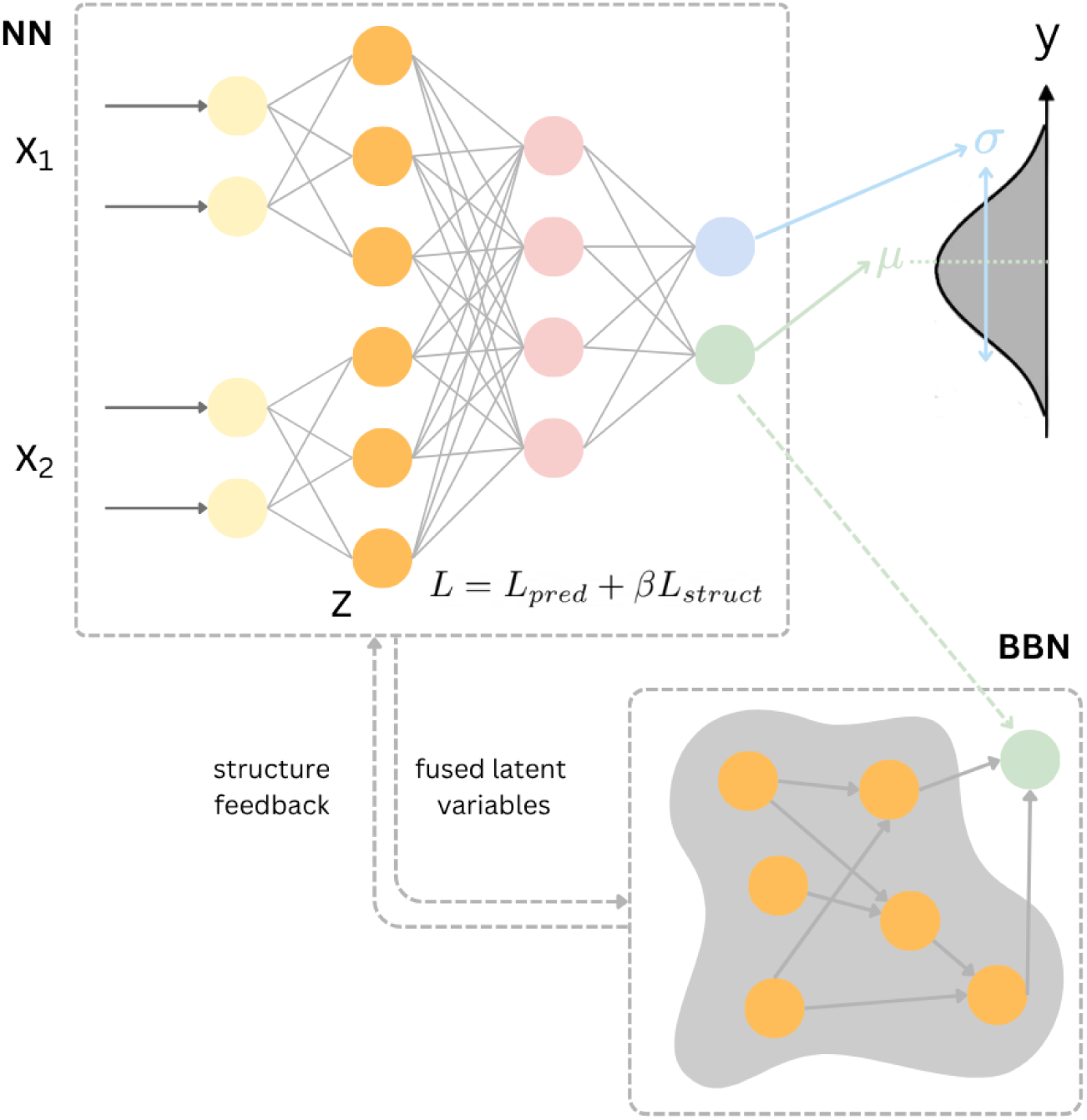
Schematic overview of the hybrid neural-Bayesian belief network framework. The cross-view latent **z** is the shared interface: it feeds the family-specific uncertainty output head (prediction branch) and the structural module (regularization branch) simultaneously.

#### DAG learner

The DAG over latent variables is learned by BIC-scored greedy hill-climbing (pgmpy HillClimbSearch [28]) applied to discretized latents. Continuous latent dimensions are discretized into four equal-frequency bins before structure search; the bin count was not varied, and the recovered DAG topology may be sensitive to this choice. A hard blacklist prevents the target node from appearing as a parent of any latent, enforcing the causal direction **z** → *y*. The DAG is refitted every Δ = 5 epochs and its edges are passed to the SN to update the active edge MLP set. Between DAG refits, the SN remains active and uses the most recently learned edge set. The first DAG fit occurs at epoch 0.

#### Structure Network

The SN provides a differentiable proxy for the DAG edge conditional distributions. For each directed edge (*i, j*) in the current DAG, it maintains a small MLP that predicts latent coordinate *z_j_* from latent coordinate *z_i_* via mean squared error (MSE), acting as a Gaussian surrogate for *P* (*z_j_* | *z_i_*). The structural loss is the mean MSE across all active edges:

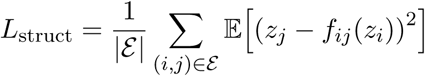

where *f_ij_* is a two-layer MLP 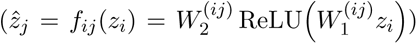 for each directed edge (*i, j*).

Edges involving the target node are excluded from SN training; the target enters only via the DAG structure constraint. The SN is updated in a separate gradient step each epoch after the BNN backward pass, using the detached latents **z** to avoid computation graph conflicts.

#### Training procedure

Each epoch alternates among three updates: (1) the neural model is updated on the combined objective *L* = *L*_pred_ + *βL*_struct_, where *L*_struct_ is computed from the current SN under the most recently fitted DAG; (2) the SN is updated on *L*_struct_ using detached latents; (3) every Δ epochs, the DAG is refit by hill-climbing and the resulting edge set is passed to the SN for subsequent epochs. When *β* = 0, the SN and DAG updates are skipped entirely, giving a pure neural baseline.

### 3.5 Experimental Design

The full GBM experiment is run as a complete matrix over:

- model family
- data view
- structural weight *β*
- cross-validation fold

For the primary predictive comparison, models are compared on the post-hoc selected primary view `t1gd_flair_mrna` and the best-performing *β* is selected per model from the observed sweep, avoiding any prior assumption about the optimal structural weight.

### 3.6 Implementation Details

All experiments used five-fold stratified cross-validation (StratifiedKFold, *k* = 5, random_state=42).

Models were optimised with Adam (learning rate 10^−3^, weight decay 10^−4^, gradient clipping at norm 1.0) for 400 epochs with no early stopping. Dropout rate was 0.3 throughout. Monte Carlo uncertainty estimation used *T* = 100 stochastic forward passes at test time for `mc_dropout` and `mc_dropout_ll`. More generally, the cross-validation protocol, optimisation settings, epoch budget, dropout rate, Monte Carlo sample count, and architecture-size rules were treated as fixed implementation settings chosen from preliminary pilot runs and then held constant across datasets, model families, and view comparisons for methodological consistency and controlled comparison rather than as claims of per-dataset optimality. No systematic hyperparameter search was performed.

### 3.7 Analysis Overview

The empirical evaluation is organized into the following analysis blocks:

- predictive performance: accuracy, macro F1, and AUC on the primary view `t1gd_flair_mrna`
- uncertainty: calibration, overconfidence, and epistemic uncertainty
- structural component analysis: *β* sensitivity and neural-only vs BBN-regularized comparison
- data-view analysis: per-view summaries, leave-one-view-out contribution, and exploratory paired comparisons against imaging baselines

This paper treats the experimental sweep as a comparative full-factor experiment rather than a single best-model report. Fold-level metrics are used to compute mean ± standard deviation. Paired fold-level comparisons are included as descriptive diagnostics alongside mean differences; given five folds and uncorrected multiple comparisons, all statistical results are treated as exploratory rather than confirmatory.

## 4 Results

### 4.1 Predictive Performance

The primary predictive analysis uses the `t1gd_flair_mrna` view, which was identified as the strongest multimodal combination in the view analysis (Section 4.5). Simple baselines on this view reached mean accuracy of 0.451 (random forest), 0.372 (MLP), and 0.363 (SVC-RBF), evaluated under the same five-fold CV and feature selection pipeline. At *β* = 0 (pure neural, no BBN), model-family mean accuracy ranged from 0.299 (bayesian) to 0.460 (nn), with AUC spanning 0.454–0.611. With structural regularization active, best-*β >* 0 configurations reached 0.468 for nn (*β* = 0.1), 0.467 for ll (*β* = 2.0), 0.468 for `mc_dropout` (*β* = 0.5), 0.460 for `mc_dropout_ll` (*β* = 2.0), and 0.331 for bayesian (*β* = 0.1). All four regularized non-Bayesian families exceeded the random-forest baseline on this view, while bayesian underperformed the other families by a wide margin. Table 3 shows the full comparison.

**Table 3:** Predictive performance on the primary view `t1gd_flair_mrna`. Simple baselines (top block) use the same five-fold CV and feature selection pipeline. Model families (bottom block) are shown at *β* = 0 (pure neural, no BBN) and at the best observed *β >* 0 (BBN active).

| Model | $\beta$ | Accuracy | Macro F1 | AUC |
| --- | --- | --- | --- | --- |
| *Simple baselines* |  |  |  |  |
| `random_forest` |  | 0.451 | 0.457 | 0.618 |
| `mlp` |  | 0.372 | 0.268 | 0.537 |
| `svc_rbf` |  | 0.363 | 0.358 | 0.514 |
| *BBN families* |  |  |  |  |
| `nn` | 0.0 | 0.460 | 0.456 | 0.611 |
|  | 0.1 | 0.468 | 0.462 | 0.613 |
| `l1` | 0.0 | 0.428 | 0.424 | 0.577 |
|  | 2.0 | 0.467 | 0.465 | 0.589 |
| `mc_dropout` | 0.0 | 0.444 | 0.441 | 0.603 |
|  | 0.5 | 0.468 | 0.464 | 0.608 |
| `mc_dropout_l1` | 0.0 | 0.411 | 0.406 | 0.580 |
|  | 2.0 | 0.460 | 0.456 | 0.587 |
| `bayesian` | 0.0 | 0.299 | 0.157 | 0.454 |
|  | 0.1 | 0.331 | 0.166 | 0.472 |

Pairwise fold-level comparisons did not show statistically reliable separation between model families, consistent with the high fold-to-fold variability expected from a small cohort.

### 4.2 Uncertainty

Uncertainty analysis on the primary view `t1gd_flair_mrna` showed a consistent pattern across model families. For the four non-Bayesian families, the best-ECE configuration occurred at *β* = 2.0. Within this group, ll yielded the lowest ECE (0.202 ± 0.062; overconfidence 0.545 ± 0.020), followed by `mc_dropout_ll` (0.214 ± 0.042; overconfidence 0.523 ± 0.024) and `mc_dropout` (0.244 ± 0.088; overconfidence 0.550 ± 0.030), while nn remained the most overconfident (ECE 0.269 ± 0.090; overconfidence 0.607 ± 0.057). The bayesian family at *β* = 0 achieved the lowest ECE overall (0.122 ± 0.031; overconfidence 0.388 ± 0.017) without structural regularization, but reliability-bin inspection showed that this result coexisted with a much narrower occupied confidence range, so it should not be interpreted as unambiguously superior calibration. The summary is shown in Figure 2.

**Figure 2:**
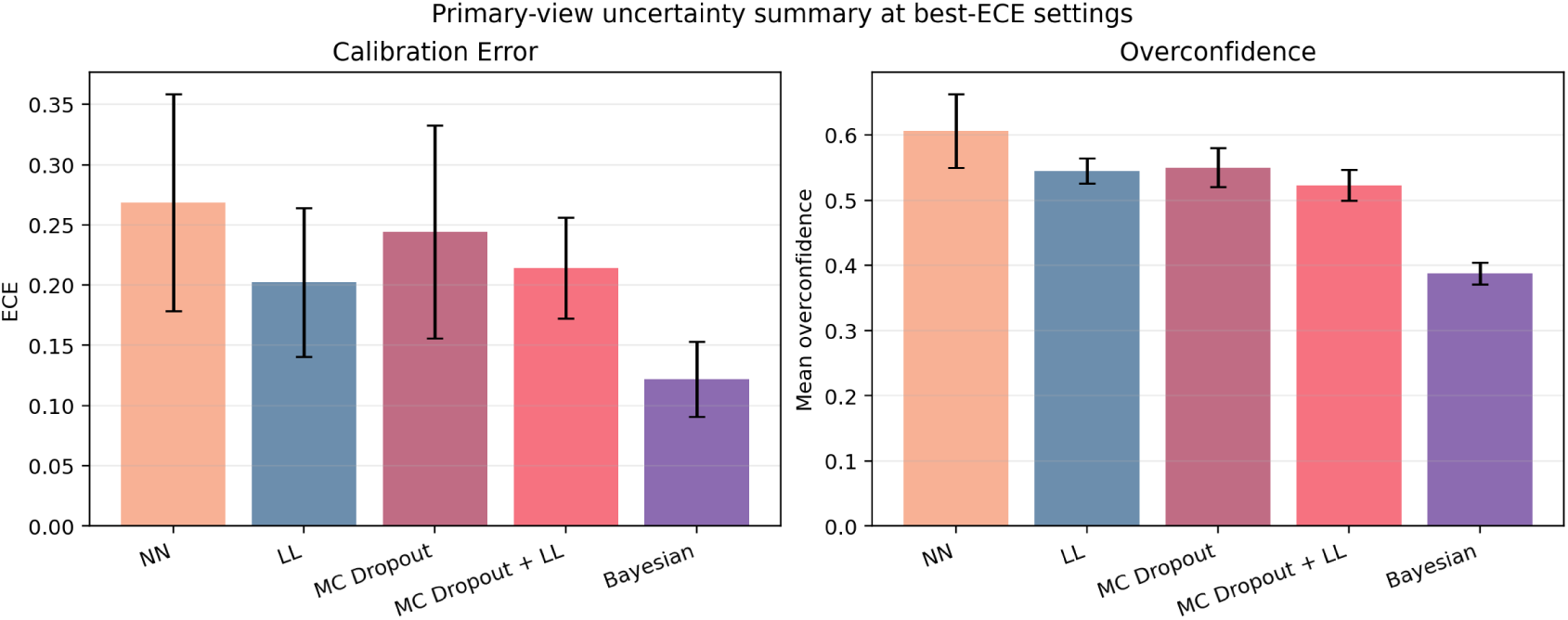
Uncertainty summary on the primary view `t1gd_flair_mrna` at the best-ECE configuration selected for each model family. bayesian achieves the lowest ECE overall, ll is the strongest non-Bayesian calibration result, and nn remains the most overconfident. Error bars show fold-level standard deviations.

Pairwise ECE comparisons showed that ll, `mc_dropout`, and `mc_dropout_ll` all outperformed nn on calibration error, consistent with the expectation that uncertainty-aware model families produce better-distributed confidence estimates than a deterministic baseline.

Reliability-bin inspection sharpens this picture. For the nn baseline, predictions concentrated in high-confidence bins yet empirical accuracy in those bins remained substantially lower than predicted confidence, showing genuine overconfidence. By contrast, ll stays closer to the diagonal, while the Bayesian panel reveals that its low ECE coexists with a much narrower occupied confidence range. These patterns are shown in Figure 3.

**Figure 3:**
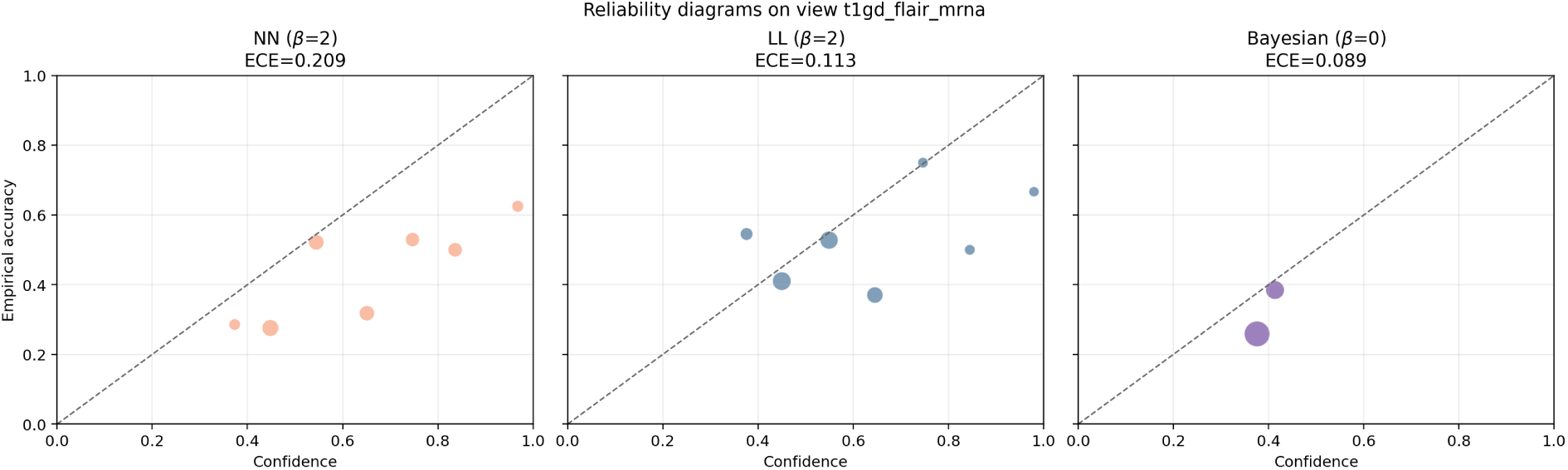
Reliability diagrams on the primary view `t1gd_flair_mrna` for the deterministic baseline (nn), the best-calibrated non-Bayesian family (ll), and the Bayesian family. Point size reflects the number of samples in each confidence bin. ll stays closer to the diagonal than nn, while the Bayesian panel shows that its low ECE is accompanied by a much narrower occupied confidence range.

Epistemic uncertainty, computed as the mean variance of predicted class probabilities across stochastic forward passes, also differed clearly across the stochastic model families on the primary view `t1gd_flair_mrna` (Figure 4). In this comparison, stochasticity arises from each family’s own uncertainty mechanism: output sampling for ll, test-time dropout for `mc_dropout`, both mechanisms for `mc_dropout_ll`, and posterior weight sampling for bayesian. At the selected configurations, bayesian showed the largest epistemic signal (0.087 ± 0.004), followed by `mc_dropout_ll` (0.025 ± 0.003), `mc_dropout` (0.018 ± 0.003), and ll (0.011 ± 0.001). These values indicate that the Bayesian family expresses substantially greater predictive variability across repeated stochastic evaluations than the heuristic families, with the MC variants showing intermediate levels of predictive uncertainty.

**Figure 4:**
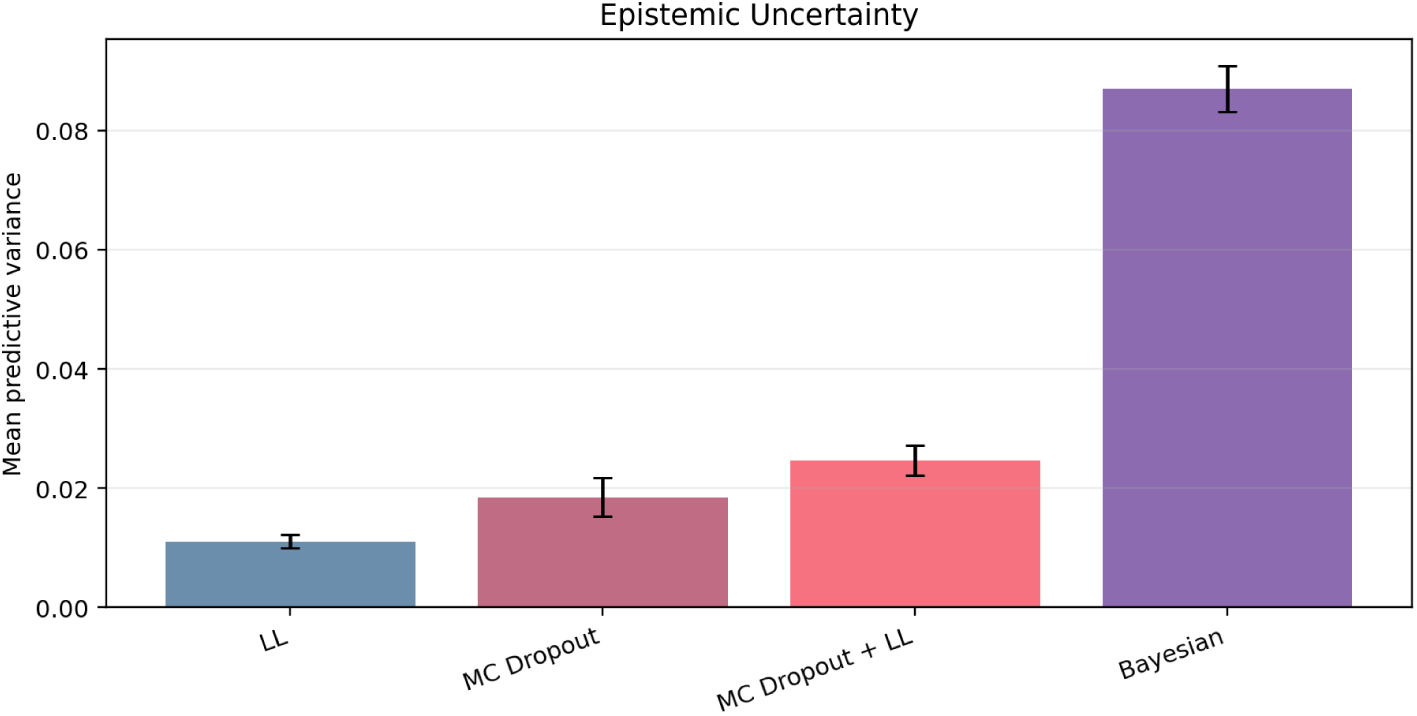
Variance-based epistemic uncertainty on the primary view `t1gd_flair_mrna` for the stochastic model families at the selected configurations. Values show the mean variance of predicted class probabilities across stochastic forward passes; error bars denote fold-level standard deviations.

### 4.3 Structural Weight Sensitivity

Structural-weight effects on the primary view `t1gd_flair_mrna` showed a model-dependent pattern across the four non-Bayesian families. Best-accuracy configurations occurred at *β* = 0.1 for nn (0.468 ± 0.069), *β* = 0.5 for `mc_dropout` (0.468 ± 0.072), and *β* = 2.0 for both ll (0.467 ± 0.102) and `mc_dropout_ll` (0.460 ± 0.104). Relative to the *β* = 0 baseline, structural regularization produced modest accuracy gains for all four non-Bayesian families: +0.008 for nn, +0.024 for `mc_dropout`, +0.039 for ll, and +0.049 for `mc_dropout_ll`. The bayesian family also improved from 0.299 ± 0.062 at *β* = 0 to 0.331 ± 0.033 at *β* = 0.1, but remained well below the other families in predictive accuracy.

Calibration followed a different pattern. For the four non-Bayesian families, the best-ECE configuration occurred at *β* = 2.0: ll (0.202 ± 0.062), `mc_dropout_ll` (0.214 ± 0.042), `mc_dropout` (0.244 ± 0.088), and nn (0.269 ± 0.090), which wereall below their respective *β* = 0 baselines (ll: 0.341 ± 0.084; `mc_dropout_ll`: 0.272 ± 0.063; `mc_dropout`: 0.269 ± 0.056; nn: 0.371 ± 0.070). The bayesian family was the exception: its lowest ECE occurred at *β* = 0.0. The accuracy cost of moving from the best-accuracy *β* to *β* = 2.0 ranged from negligible (`mc_dropout_ll`: 0.000; ll: 0.000) to moderate (nn: −0.048; `mc_dropout`: −0.073). The accuracy–calibration trade-off therefore remains clear for the non-Bayesian families, although the precise best-*β* operating point is model-dependent.

As a component-level diagnostic, we compared neural and graphical prediction accuracy at the best-accuracy configuration per model on the primary view (Table 4). The neural predictor consistently outperformed the graphical component across all five families, with differences ranging from 0.089 (`mc_dropout_ll`) to 0.251 (bayesian). The graphical component provides supplementary structure but does not match the neural predictor as a standalone classifier on this view.

**Table 4:** Neural-versus-graphical accuracy comparison on the primary view `t1gd_flair_mrna` at the best-accuracy configuration per model. The neural predictor consistently outperforms the graphical (BBN) component.

| Model | $\beta$ | Neural acc | Graphical acc | Difference |
| --- | --- | --- | --- | --- |
| `nn` | 0.1 | 0.468 | 0.338 | 0.130 |
| `l1` | 2.0 | 0.467 | 0.371 | 0.096 |
| `mc_dropout` | 0.5 | 0.468 | 0.322 | 0.146 |
| `mc_dropout_l1` | 2.0 | 0.460 | 0.371 | 0.089 |
| `bayesian` | 0.1 | 0.331 | 0.080 | 0.251 |

**Table 5:** Mean accuracy across all 15 view combinations (best *β* per model selected on `t1gd_flair_mrna`: nn *β*=0.1, ll *β*=2.0, `mc_dropout` *β*=0.5, `mc_dropout_ll` *β*=2.0, bayesian *β*=0.1). Bold values are within 0.01 of the column maximum. Views containing cna usually underperform their cna-free counterparts, with the clearest and most consistent degradation in the four non-Bayesian families.

| View | nn | l1 | mc_dropout | mc_dropout_l1 | bayesian |
| --- | --- | --- | --- | --- | --- |
| *Single-view* |  |  |  |  |  |
| `cna` | 0.264 | 0.201 | 0.233 | 0.201 | 0.333 |
| `flair` | 0.380 | 0.403 | 0.418 | 0.388 | 0.336 |
| `mrna` | 0.379 | 0.411 | 0.371 | 0.387 | 0.315 |
| `t1gd` | 0.410 | 0.350 | 0.403 | 0.335 | 0.328 |
| *Two-view* |  |  |  |  |  |
| `flair_cna` | 0.340 | 0.341 | 0.309 | 0.333 | 0.333 |
| `flair_mrna` | 0.315 | 0.372 | 0.364 | 0.348 | 0.299 |
| `mrna_cna` | 0.354 | 0.387 | 0.370 | 0.353 | 0.319 |
| `t1gd_cna` | 0.318 | 0.326 | 0.325 | 0.325 | 0.333 |
| `t1gd_flair` | 0.419 | 0.381 | 0.441 | 0.374 | 0.305 |
| `t1gd_mrna` | 0.355 | 0.396 | 0.355 | 0.387 | 0.315 |
| *Three-view* |  |  |  |  |  |
| `flair_mrna_cna` | 0.370 | 0.387 | 0.387 | 0.412 | **0.353** |
| `t1gd_flair_cna` | 0.348 | 0.380 | 0.410 | 0.387 | 0.341 |
| `t1gd_flair_mrna` | **0.468** | **0.467** | **0.468** | **0.460** | 0.331 |
| `t1gd_mrna_cna` | 0.354 | 0.311 | 0.311 | 0.303 | **0.345** |
| *Four-view* |  |  |  |  |  |
| `t1gd_flair_mrna_cna` | 0.319 | 0.353 | 0.327 | 0.353 | 0.319 |

### 4.4 Learned DAG Structure

Structural analysis of the learned BBN graphs provides a qualitative picture of the regularization component’s behavior. The DAG learner operates on the output of the cross-view fusion layer, a cross_latent_dim-dimensional mixed representation of all input modalities, so individual latent nodes cannot be attributed to specific views. The BIC hill-climber produces sparse graphs in which only a subset of latent dimensions participate in any edge; the active subgraph averages roughly 15 edges over 1516 nodes for `t1gd_flair_mrna`, with the remaining latent dimensions isolated.

DAG structure is poorly reproducible across cross-validation folds. Figure 5a shows mean pair-wise Jaccard similarity between fold-final graphs for all five model families across *β* ∈ {0.1, 0.5, 1.0, 2.0}. For the primary view, the mean Jaccard similarity ranged is 0.00–0.19, with the Bayesian family flat at zero across all *β* values (indicating no reproducible edge overlap across folds) and the non-Bayesian families remaining low and variable. There is no consistent trend across *β* or model family; stronger regularization does not produce more reproducible structure.

**Figure 5:**
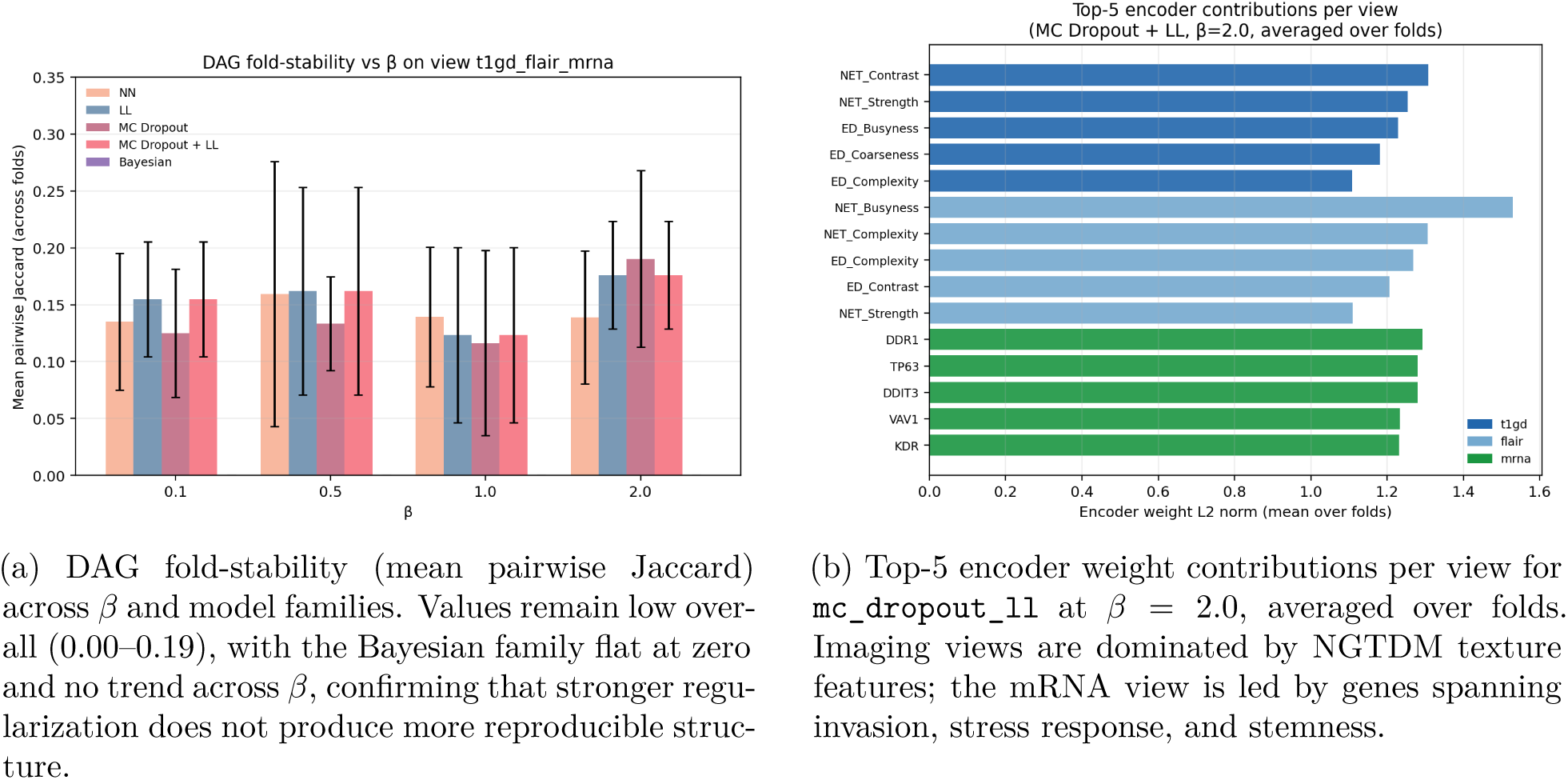
DAG structure analysis on the primary view `t1gd_flair_mrna`. **(a)** Graph reproducibility is uniformly low across all configurations. **(b)** Despite DAG instability, encoder feature rankings are consistent across folds, indicating that the latent space encodes biologically coherent input signal.

Although the DAG operates on a mixed latent space, the encoder weight matrices connect those latents back to input features. Figure 5b shows the top-5 encoder weight contributions per view for `mc_dropout_ll` at *β* = 2.0, averaged over all five folds. The imaging views (t1gd and flair) are dominated by NGTDM texture features from the necrotic tumor (NET) and edematous/invaded tissue (ED) sub-regions. The mRNA view is led by *DDR1*, *TP63*, *DDIT3*, *VAV1*, and *KDR*. The consistency of this ranking across folds suggests that the latent representations, despite being structured by an unstable DAG, encode signal from coherent input features.

### 4.5 Data-View Contribution

The data-view analysis showed that view choice mattered at least as much as model family. Table 5 reports mean accuracy for all 15 view combinations, using the best *β* per model selected on the primary view `t1gd_flair_mrna` (nn: *β*=0.1; ll: *β*=2.0; `mc_dropout`: *β*=0.5; `mc_dropout_ll`: *β*=2.0; bayesian: *β*=0.1). For the four non-Bayesian families, the strongest results were concentrated in the three-view combination `t1gd_flair_mrna`, which achieved the highest mean accuracy (nn: 0.468; ll: 0.467; `mc_dropout`: 0.468; `mc_dropout_ll`: 0.460). Imaging-only baselines t1gd_flair and flair were also competitive for these families, while the full four-view setting `t1gd_flair_mrna_cna` remained clearly weaker than the best selective-fusion alternatives. The bayesian family was the exception: its strongest views were the cna-containing three-view combinations flair_mrna_cna (0.353) and t1gd_mrna_cna (0.345).

Leave-one-view-out (LOVO) analysis tested which modalities contributed positively to the full four-view setting `t1gd_flair_mrna_cna`. We defined contribution as accuracy(full-view) − accuracy(full-view minus one view), so negative values indicate that removing a view *improves* accuracy. Dropping cna produced the largest and most consistent improvement (at *β* = 0.0): −0.141 (nn), −0.117 (ll), −0.167 (`mc_dropout`), and −0.067 (`mc_dropout_ll`). At this level, dropping mrna or t1gd also improved accuracy for most families; however, this does not mean those modalities are independently harmful. To test this, we computed marginal contributions across all 15 view subsets of the four modalities (1-view through 4-view contexts). As a single view, cna alone achieved mean accuracy of 0.264 (nn), 0.201 (ll), 0.233 (`mc_dropout`), and 0.201 (`mc_dropout_ll`), which were well below every other single-view baseline and near or below chance for a three-class problem. Across multi-view contexts, cna showed a negative marginal contribution in the large majority of contexts in which it appeared (mean Δ accuracy: −0.058 to −0.067 averaged across all contexts). Isolated exceptions exist at the 2-view level (mrna_cna marginally exceeded mrna for `mc_dropout`: 0.378 vs 0.370) and the 3-view level without T1Gd (flair_mrna_cna exceeded flair_mrna for nn, ll, and `mc_dropout`), but these gains were small and inconsistent across model families. By contrast, t1gd and mrna had positive contributions in cna-free contexts and near-zero or negative contributions only in cna-containing contexts. flair was consistently positive across all contexts. The apparent degradation of t1gd and mrna at the 4-view level is therefore a consequence of cna lowering the performance of the whole combination, not an independent property of those modalities. Figure 6 shows the 4-view LOVO breakdown.

**Figure 6:**
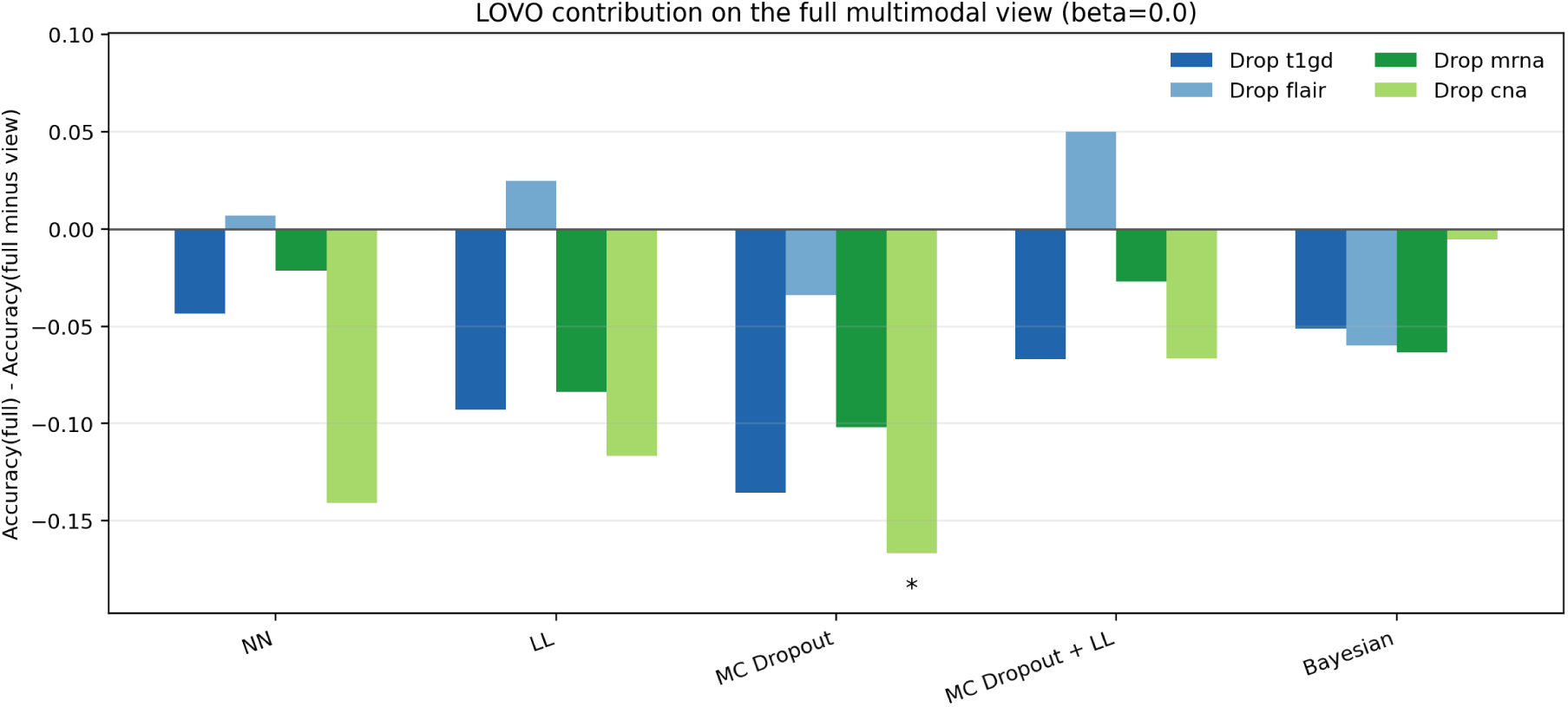
Leave-one-view-out contribution from the full four-view baseline `t1gd_flair_mrna_cna` at *β* = 0.0. Negative values indicate that removing a view improves accuracy. cna shows the largest and most consistent degradation effect; flair is the only modality whose removal does not reliably help.

To test whether multimodal views improved on imaging-only baselines, we computed paired fold-level differences against each model’s better imaging baseline (t1gd or flair) at *β* = 0.0, reporting *p*-values as descriptive alongside mean differences. Positive effects remained rare. The clearest was for nn, where `t1gd_flair_mrna` showed a mean improvement of 0.065 over flair (*p* = 0.033). Negative effects were more common and generally larger, especially for views containing cna. Table 6 therefore reports one row per model family using that model’s strongest degradation relative to its imaging baseline under this *β* = 0.0 view-isolation analysis.

**Table 6:** Paired comparisons at *β* = 0.0 against the best imaging baseline chosen separately for each model. One row is shown per model family, using that family’s strongest degradation relative to its imaging baseline. Positive values indicate improvement over the imaging baseline; negative values indicate degradation.

| Model | View | Baseline | Mean ± SD | $p$ |
| --- | --- | --- | --- | --- |
| nn | t1gd_mrna_cna | flair | $-0.083 \pm 0.130$ | 0.226 |
| ll | t1gd_cna | flair | $-0.132 \pm 0.118$ | 0.066 |
| mc_dropout | t1gd_flair_mrna_cna | flair | $-0.156 \pm 0.161$ | 0.096 |
| mc_dropout_ll | t1gd_cna | flair | $-0.147 \pm 0.117$ | 0.049 |
| bayesian | t1gd_flair_mrna_cna | t1gd | $-0.035 \pm 0.037$ | 0.107 |

Taken together, the view analyses support a clear finding: cna consistently degrades performance across the four non-Bayesian model families, whereas the bayesian family’s strongest views include cna. flair is the most reliable contributor; t1gd and mrna carry positive signal in imaging-only and genomic combinations but are suppressed when paired with cna. For the four non-Bayesian model families, selective fusion, specifically `t1gd_flair_mrna`, outperforms both the full four-view baseline and imaging-only alternatives, confirming that view selection matters at least as much as model choice in this cohort.

### 4.6 HAR Benchmark

To assess whether the observed GBM performance ceiling reflects a general architectural limitation or instead the cohort’s size and signal quality, we applied the same five-family framework to a human activity recognition (HAR) dataset with many more samples, six activity classes, and inertial-sensor views. A complete five-fold experiment was conducted on two HAR views: a time-domain sensor view (`t_acc_t_gyro`, post-hoc designated primary) and a frequency-domain sensor view (f_acc_f_gyro).

On the primary view `t_acc_t_gyro`, the four non-Bayesian families achieved near-ceiling mean accuracy of 0.940–0.941 at their best positive-*β* settings, compared with 0.468 for the best GBM configuration on `t1gd_flair_mrna`. The bayesian family reached 0.781 at *β* = 2.0, remaining clearly below the other HAR families but substantially above chance. The corresponding best accuracy on the frequency-domain view f_acc_f_gyro remained high for the four non-Bayesian families (0.888–0.891) and was 0.743 for bayesian. These results indicate that the framework can achieve strong predictive performance when the underlying signal is strong, suggesting that the GBM performance ceiling is more likely driven by data limitations than by an architectural failure. The structural-regularization pattern also extends to HAR, but the gains remain modest. Best positive-*β* settings fall at *β* = 0.5 for nn and `mc_dropout`, at *β* = 1.0 for ll and `mc_dropout_ll`, and at *β* = 2.0 for bayesian. Relative to the *β* = 0 baseline, the primary-view accuracy gain is +0.009 for nn, +0.011 for ll, +0.008 for `mc_dropout`, +0.011 for `mc_dropout_ll`, and +0.006 for bayesian (Table 7). As in GBM, BBN regularization helps directionally, but the effect size is small relative to the overall performance level.

**Table 7:** HAR results on the primary view `t_acc_t_gyro` at *β* = 0 (no BBN) and at the best observed *β >* 0 configuration for each model family. Values are mean ± SD over five folds.

| Model | $\beta$ | Accuracy |
| --- | --- | --- |
| **nn** | 0.0 | $0.932 \pm 0.009$ |
| | 0.5 | $0.940 \pm 0.007$ |
| **ll** | 0.0 | $0.930 \pm 0.009$ |
| | 1.0 | $0.941 \pm 0.007$ |
| **mc_dropout** | 0.0 | $0.933 \pm 0.008$ |
| | 0.5 | $0.941 \pm 0.008$ |
| **mc_dropout_ll** | 0.0 | $0.930 \pm 0.008$ |
| | 1.0 | $0.941 \pm 0.007$ |
| **bayesian** | 0.0 | $0.775 \pm 0.023$ |
| | 2.0 | $0.781 \pm 0.042$ |

The uncertainty story differs more sharply from GBM. On HAR, the lowest-ECE configuration for the four non-Bayesian families occurs at *β* = 0, not at higher structural weights, indicating that these models are already well-calibrated on the easier, larger-sample task before regularization is applied. Overconfidence remains highest for the deterministic and likelihood-style families (nn: 0.710 ± 0.021; ll: 0.707 ± 0.018), lower for the MC variants (`mc_dropout`: 0.648 ± 0.017; `mc_dropout_ll`: 0.646 ± 0.013), and lowest for bayesian (0.573 ± 0.032 at *β* = 0.5). As in GBM, however, lower overconfidence does not imply the strongest predictive performance: Bayesian remains the weakest HAR family in accuracy despite being the least overconfident. Uncertainty summaries, structural-sensitivity summaries, and DAG diagnostics for the HAR benchmark are reported in Appendix A.

Together, the HAR results support the conclusion that the GBM performance ceiling (∼0.46) reflects data limitations rather than an architectural one. The same framework reaches *>* 0.90 accuracy when signal is adequate, and the directionally positive effect of structural regularization appears across both data regimes even though the family-specific best positive-*β* settings differ.

## 5 Discussion

The results support interpreting the hybrid framework primarily as an uncertainty-aware multi-modal predictive model, not as a structure-discovery system. Deterministic and heuristic uncertainty-aware families were generally more competitive on predictive accuracy than the fully probabilistic Bayesian variant. The Bayesian branch is most useful here not as the strongest predictive model, but as a contrasting uncertainty formulation that achieved the lowest ECE under a narrower occupied confidence range, highlighting the trade-offs among predictive strength, calibration, and epistemic uncertainty in a small-sample multimodal setting [5–7].

The multimodal findings require careful interpretation. The LOVO analysis shows that flair is the only view whose removal from the four-view combination does not consistently improve accuracy, and it is among the most informative single modalities (best single-view accuracy for two of four non-Bayesian model families). Removing cna, mrna, or t1gd from the four-view combination each improves accuracy, with cna having the largest and most consistent effect. However, the complete marginal contribution analysis across all 15 view subsets (Section 4.5) shows that this degradation is not symmetric: cna is the only modality that hurts performance in the large majority of contexts in which it appears (with isolated exceptions at the 2- and 3-view level for some families), while t1gd and mrna are positive contributors in cna-free combinations and appear harmful only when cna is present. The 4-view LOVO result for t1gd and mrna therefore reflects contamination by the cna combination rather than an independent degradation effect. For the four non-Bayesian families, the best multimodal result comes from selectively combining t1gd, flair, and mrna, a three-view subset that outperforms the full four-view setting. The bayesian family is the exception: its strongest results shift toward cna-containing three-view combinations, consistent with its notably higher single-view CNA accuracy (0.333) relative to all other families (0.201–0.264). One possible explanation is that posterior weight uncertainty provides a form of noise regularization that partially attenuates CNA’s low signal quality, though this mechanism was not directly tested. The view analysis therefore separates genuinely informative multimodal combinations from those that add complexity without clear predictive benefit, consistent with the broader radiogenomic literature’s emphasis on heterogeneity across imaging-molecular combinations rather than automatic gains from fusion alone [4].

The consistent degradation from cna is biologically interpretable. GBM copy number alterations derived from bulk SNP array profiling are subject to substantial intratumoral spatial heterogeneity: common focal events vary across biopsy sites within the same tumor, meaning a single-site profile provides an incomplete genomic snapshot [29, 30]. The most prevalent CNA events in IDH-wildtype GBM, including EGFR amplification, present in approximately 40% of cases, do not independently predict overall survival in multivariable analyses within this subtype [31, 32]. Prior comparative analyses across TCGA cohorts have also shown that mRNA expression features outperform copy number features for survival prediction, because expression profiles capture transcriptional programs more directly than the upstream CNA events that partially drive them [33]. Together, these considerations offer a plausible biological explanation for the CNA degradation, though the causal mechanisms were not directly tested and the interpretation remains speculative.

The uncertainty findings make a clear practical point. On the primary view `t1gd_flair_mrna`, the deterministic nn baseline is the most overconfident: it assigns high confidence to predictions that are frequently wrong, which is a real clinical concern. The uncertainty-aware non-Bayesian families (ll, `mc_dropout`, `mc_dropout_ll`) all produce better-calibrated estimates, with ll yielding the strongest non-Bayesian calibration result. The bayesian family achieved the lowest ECE overall but with a much narrower occupied confidence range, which limits the scope of that advantage. Beyond ECE, the stochastic families differ substantially in epistemic uncertainty, the mean variance of predicted class probabilities across repeated stochastic evaluations. bayesian expressed the largest epistemic signal (0.087 ± 0.004), more than three times that of `mc_dropout_ll` (0.025 ± 0.003), `mc_dropout` (0.018 ± 0.003), and ll (0.011 ± 0.001). This high predictive variance identifies cases where the model’s output is genuinely unstable across evaluations, independently of calibration, and may be clinically useful for flagging predictions that warrant additional scrutiny. The overall trade-off is therefore three-dimensional: nn and `mc_dropout` achieve the highest accuracy, ll offers the strongest non-Bayesian calibration, and bayesian combines the lowest ECE with the highest epistemic uncertainty expression at substantially weaker predictive accuracy. None of these properties uniformly dominates, which is a recurring feature of uncertainty-aware medical AI in small-cohort settings [7, 13].

The structural regularization component produced a consistent but model-dependent pattern. Across the four non-Bayesian families, activating the BBN regularizer improved predictive accuracy relative to *β*=0, but the best operating point differed by family: nn at *β*=0.1, `mc_dropout` at *β*=0.5, and both ll and `mc_dropout_ll` at *β*=2.0. Calibration showed a cleaner pattern: *β*=2.0 achieved lower ECE than *β*=0 for every non-Bayesian family, indicating that stronger structural regularization systematically smooths confidence estimates regardless of which *β* is optimal for accuracy. The bayesian family was the exception: its lowest ECE occurred at *β*=0.0, consistent with the interpretation that posterior weight uncertainty already imposes substantial regularization, leaving little for the DAG component to contribute and instead compressing the occupied confidence range. The neural-versus-graphical comparison (Table 4) makes the BBN’s role explicit: at best-*β* configurations, the graphical component achieved 0.080–0.371 accuracy across all five families, consistently below the neural predictor (0.331–0.468), confirming that the BBN contributes to the training objective without acting as a competitive standalone classifier. These patterns are observational and should not be interpreted as recovered dependencies between modalities or biological variables; the BBN functions here as a learned latent regularizer rather than a structure-discovery mechanism. Which operating point is preferable depends on clinical priority: lower *β* favours accuracy, higher *β* favours calibration.

The learned DAG structures (Section 4.4) are sparse and poorly reproducible across folds. Mean pairwise Jaccard similarity between fold-final graphs ranged from 0.00 to 0.19 on the primary view, with the bayesian family flat at zero across all *β* values and the non-Bayesian families remaining low and variable. Stronger regularization did not produce more reproducible structure: there was no consistent trend in fold-stability across *β* or model family. This instability is not unexpected at *n* = 134: hill-climbing BIC search over a discretized latent space is sensitive to the fold composition, and the graph is learned on a mixed cross-view representation rather than view-separated latents, limiting biological attribution of individual edges. The contribution of the BBN component is therefore at the regularization level rather than the structure-recovery level.

Despite the DAG instability, the encoder weight matrices connecting input features to the latent space are consistent across folds. The imaging views (t1gd and flair) are dominated by NGTDM texture features from the necrotic tumor (NET) and edematous/invaded tissue (ED) subregions, whose prominence may reflect the biologically distinct character of GBM’s necrotic core and peritumoral edema. The mRNA view is led by *DDR1*, *TP63*, *DDIT3*, *VAV1*, and *KDR*, which are genes spanning collagen-mediated invasion, stress-state transcriptional programming, and tumour microenvironment signalling [34–36], though their individual roles in survival prediction were not validated. The fold-stability of this feature ranking, even when the DAG itself is not stable, suggests that the encoder representations capture a consistent signal, and that the BBN regularizer operates on a meaningful latent space even if it cannot recover a reliable graph structure at this sample size. The HAR benchmark confirms that the GBM performance ceiling reflects data limitations rather than an architectural one. The four non-Bayesian families reached near-ceiling accuracy (0.940–0.941) at best positive-*β* settings, compared with 0.468 for the best GBM configuration; the bayesian family reached 0.781, remaining the weakest family but far above its near-random GBM behaviour. Structural regularization extended to HAR with consistent directional gains (+0.006– +0.011 over *β*=0), similar in character but smaller in relative magnitude to the GBM gains. The calibration story shifted: for the four non-Bayesian families, the lowest ECE occurred at *β*=0 rather than at higher structural weights, the reverse of the GBM finding. This is interpretable by task difficulty: on a larger, higher-signal dataset models are already well-calibrated without regularization, whereas the calibration benefit of higher *β* on GBM may reflect structural regularization’s role in smoothing confidence estimates when signal is weak and the sample is small. DAG stability remained uniformly low on HAR as well, reinforcing the conclusion that the BBN component operates as a mild latent regularizer rather than a reliable structure-recovery mechanism regardless of data regime.

Several limitations bear on interpretation. The primary cohort comprises 134 patients from TCGA, which is small relative to the number of features and model parameters involved. The survival outcome was discretized into three classes using fixed thresholds derived from the cohort distribution, and results may be sensitive to this choice. Cross-validation with five folds provides stable average estimates but leaves fold-level comparisons underpowered; all statistical contrasts should be treated as exploratory. Calibration estimates carry high variance when computed over test sets of roughly 25–30 patients per fold, and the relative ECE rankings across families should not be read as precise or stable quantitative distinctions.

No systematic hyperparameter search was performed; learning rates, dropout rates, network widths, and the latent dimension scaling rule (*d*_lat_ = ⌊*d_v_/*2⌋) were fixed to values established in preliminary runs and held constant across all comparisons. Training used a fixed epoch budget without early stopping, introducing a risk of overfitting in later epochs that cannot be ruled out without validation-curve monitoring.

Within each training fold, features were selected per view by univariate Cox partial-likelihood ranking and the top-*k* retained; *k* was fixed at 10 for all views and was not optimised per view; a different selection threshold could affect view-specific results. All view encoders share the same latent dimensionality, meaning modalities with very different intrinsic dimensionality are treated as equally informative by the fusion layer, a constraint that may contribute to the consistent CNA degradation. The latent-space discretization applied before DAG structure search (four equal-frequency bins) was not varied, and the exact differences among *β>*0 configurations should not be over-interpreted as invariant optima: the learned graph depends on a hill-climbing path that is only partially controlled.

The strongest conclusion is not that the framework solved multimodal GBM prediction out-right, but that it makes a set of practically relevant trade-offs visible: selective multimodal fusion outperforms naive four-view inclusion, calibration quality and epistemic uncertainty signal differ sharply across families in ways that matter for clinical use, and structural regularization acts as a useful latent regularizer without supporting structure-discovery claims at this sample size. The HAR results situate the GBM performance ceiling as a data limitation rather than an architectural one, suggesting that the framework’s behaviour generalises when the underlying signal is adequate.

## 6 Conclusion

This paper presented a systematic evaluation of a hybrid neural network–Bayesian belief network framework for multimodal GBM survival prediction across five model families, fifteen view combinations, and five structural regularization weights. The main contribution is a structured account of how predictive accuracy, calibration quality, and epistemic uncertainty trade off across families, and how view selection determines the multimodal signal available in a small clinical cohort. On the primary view `t1gd_flair_mrna`, the non-Bayesian families outperformed the random forest baseline, though pairwise accuracy differences between families remain exploratory at this sample size.

Three findings stand out. First, CNA copy-number features consistently degraded predictive performance across most view contexts, and selective fusion excluding CNA outperformed both the full four-view baseline and imaging-only alternatives; modality choice proved at least as important as model choice, and the degradation is biologically interpretable. Second, uncertainty behaviour separated clearly across families: heuristic uncertainty-aware families achieved better calibration than the deterministic baseline, the Bayesian family reached the lowest expected calibration error but over a narrower confidence range and at substantially weaker discriminative performance, limiting the practical scope of that advantage, and epistemic uncertainty signals differed by an order of magnitude between families. These are differences with direct implications for how model outputs should be interpreted in practice. Third, BBN structural regularization produced consistent directional benefits for both accuracy and calibration without supporting structure-discovery claims; the BBN’s contribution is as a latent regularizer rather than a structure-recovery mechanism, and learned graph structures were not reproducible at this sample size.

Together, these findings position the study as a practical multimodal prediction and uncertainty analysis rather than a structure-discovery framework. The HAR benchmark supports this framing: the same framework reaches near-ceiling accuracy when the underlying signal is adequate, confirming that the GBM performance ceiling is a data limitation rather than an architectural one. The framework’s contribution is the structured visibility it provides over trade-offs among accuracy, calibration, epistemic uncertainty, and modality selection — dimensions that matter for how uncertainty-aware models should be evaluated and selected in small-cohort clinical settings.

## A HAR Benchmark: Additional Analysis

To mirror the main GBM analysis sections, Tables 8–11 summarize uncertainty behavior, structural-weight sensitivity, and DAG characteristics for the HAR primary view `t_acc_t_gyro`.

**Table 8:** HAR primary-view uncertainty summary at the best-ECE configuration for each model family. Values are mean ± SD over five folds. For the four non-Bayesian families, the lowest ECE occurs at *β* = 0, whereas Bayesian reaches its best ECE at *β* = 0.5.

| Model | $\beta_{\text{ECE}}$ | ECE | Overconfidence | Epistemic |
| --- | --- | --- | --- | --- |
| nn | 0.0 | $0.011 \pm 0.005$ | $0.710 \pm 0.021$ | $0.000 \pm 0.000$ |
| ll | 0.0 | $0.012 \pm 0.005$ | $0.707 \pm 0.018$ | $0.000 \pm 0.000$ |
| mc_dropout | 0.0 | $0.046 \pm 0.005$ | $0.648 \pm 0.017$ | $0.006 \pm 0.000$ |
| mc_dropout_ll | 0.0 | $0.049 \pm 0.008$ | $0.646 \pm 0.013$ | $0.006 \pm 0.000$ |
| bayesian | 0.5 | $0.069 \pm 0.037$ | $0.573 \pm 0.032$ | $0.037 \pm 0.004$ |

**Table 9:** HAR primary-view structural-weight sensitivity summary. ‘Acc (*β*=0)‘ is the pure-neural baseline; ‘Best acc’ is the strongest configuration over *β >* 0; ‘PGM acc’ is the graphical-component accuracy at that best-accuracy configuration. Structural gains are modest for all families, while Bayesian remains substantially below the other models despite improving slightly at *β* = 2.0.

| Model | Acc ($\beta=0$) | $\beta_{\text{best acc}}$ | Best acc | PGM acc |
| --- | --- | --- | --- | --- |
| nn | 0.932 | 0.5 | 0.940 | 0.888 |
| ll | 0.930 | 1.0 | 0.941 | 0.904 |
| mc_dropout | 0.933 | 0.5 | 0.941 | 0.890 |
| mc_dropout_ll | 0.930 | 1.0 | 0.941 | 0.904 |
| bayesian | 0.775 | 2.0 | 0.781 | 0.123 |

**Table 10:** HAR primary-view DAG fold-stability (mean pairwise Jaccard similarity) across structural-weight settings. Stability remains low overall for all model families and is especially weak for Bayesian.

| Model | $\beta = 0.1$ | $\beta = 0.5$ | $\beta = 1.0$ | $\beta = 2.0$ |
| --- | --- | --- | --- | --- |
| nn | 0.086 | 0.082 | 0.094 | 0.080 |
| ll | 0.091 | 0.097 | 0.079 | 0.099 |
| mc_dropout | 0.102 | 0.081 | 0.073 | 0.080 |
| mc_dropout_ll | 0.091 | 0.097 | 0.079 | 0.099 |
| bayesian | 0.028 | 0.028 | 0.028 | 0.028 |

**Table 11:** Representative HAR primary-view DAG structure summary at the best-accuracy configuration for each model family. The active HAR subgraphs are much denser than the GBM primary-view graphs, with nearly the full latent space participating.

| Model | $\beta$ | Rep. fold | Active nodes | Active edges | Outcome parents |
| --- | --- | --- | --- | --- | --- |
| nn | 0.5 | 2 | 51 | 112 | latent13, latent21, latent4 |
| ll | 1.0 | 4 | 51 | 120 | latent14, latent20, latent27 |
| mc_dropout | 0.5 | 2 | 51 | 116 | latent13, latent24, latent5 |
| mc_dropout_ll | 1.0 | 4 | 51 | 120 | latent14, latent20, latent27 |
| bayesian | 2.0 | 2 | 51 | 95 | latent14, latent33 |

## Ethics Statement

This study did not involve any new experiments on human participants or animals conducted by the authors. The glioblastoma analysis used previously collected, publicly available, de-identified datasets from TCGA/cBioPortal and TCIA/BraTS-TCGA-GBM, and the human activity recognition benchmark used the publicly available UCI HAR dataset. The authors analyzed these existing datasets in accordance with the terms of use of the respective data sources. No new ethical approval or informed consent was required for this secondary analysis of publicly available, de-identified data.

## Declaration of Generative AI and AI-Assisted Technologies in the Writing Process

During the preparation of this work the author(s) used Claude (Anthropic) and ChatGPT (OpenAI) in order to improve readability and language clarity. After using this tool/service, the author(s) reviewed and edited the content as needed and take(s) full responsibility for the content of the publication.

## Author Contributions

A.J.: Conceptualisation, Methodology, Software, Formal Analysis, Writing – Original Draft. V.H.: Supervision, Writing – Review & Editing.

## Data Availability

Code and processed TCGA-GBM feature datasets will be made available upon publication. Raw imaging data are available from The Cancer Imaging Archive (TCIA). Raw genomic data are available from cBioPortal.

